# Alcohol, smoking, and illicit substance use in Cameroon: unveiling related risk factors among university students in Yaoundé

**DOI:** 10.1101/2024.03.10.24304034

**Authors:** Fabrice Zobel Lekeumo Cheuyem, Michel Franck Edzamba, Adidja Amani, Tatiana Mossus

## Abstract

**Background:** Substance use, including alcohol, tobacco and illicit drugs, is a growing public health problem worldwide. There is a rapid increase in substance use among young adults in many sub-Saharan African countries. This study aimed to assess the factors associated with the use of different psychoactive substances among university students in Yaoundé.

**Methods:** A cross-sectional and analytical study was conducted from September to October 2023 at Yaoundé 1 University in Cameroon. All eligible students aged 18 years and older who gave written informed consent were included. A convenience nonprobability sampling method was used to recruit consenting students. The data collectors were medical students who were trained for 2 days and given appropriate instructions before the survey. The data collected were reviewed and checked for completeness before being entered. The data were analyzed using Statistics 4.3.1.

**Results:** A total of 191 university students were enrolled in the study. Age (*p*-value=0.002), level of study (*p*-value=0.048), and smoking status (*p*-value=0.005) of the participants were significant factors associated with alcohol on univariate analysis. Multivariate logistic regression showed that students aged 20-25 years were significantly 2.9 times more likely to drink alcohol than those aged less than 20 years (*p*-value=0.003). Students who smoke were 2.7 times more likely to drink alcohol than those who do not smoke (*p*-value=0.008). Living situation (p=0.013) and drug use status (*p*-value<0.0001) were significant factors associated with smoking on univariate analysis. On multivariate analysis, drug users were 3.2 times more likely to smoke than drug non-users (*p*-value<0.0001). Drug use was significantly associated with district of residence of consumer on univariate analysis (*p*-value=0.024). Living situation (*p*-value=0.016), faculty/school(*p*-value=0.04), and district of residence (*p*-value=0.037) were significantly associated with polysubstance use. Students living in shared accommodation were 3.8 times more likely to be polysubstance users than those living with their families (*p*-value=0.023). Almost all smokers (95.1%) reported being aware the of the psychosocial, mental and health consequences of substance use (*p*-value=0.021).

**Conclusion:** Several factors have been associated with substance use among college students. These sociodemographic factors can help to strategize and implement tailored interventions to reduce the risk of subsequent substance dependence and other harmful consequences.

## Background

Substance use, including alcohol, tobacco and illicit drugs, is a growing public health and socioeconomic problem worldwide [1]. Alcohol is a psychoactive substance with addictive properties that has been widely used in many cultures for centuries. Harmful alcohol use places a heavy burden on societies in terms of disease, social and economic costs. The use of psychoactive substances such as cannabis, amphetamines, cocaine, opioids, and non-prescribed psychoactive prescription medication causes significant health and social problems for the people who use them, and also for others in their families and communities [2]. The global burden of disease attributable to alcohol and illicit drug accounts 5.4% of the total burden of disease [3].

Study reports underscore the high likelihood of addiction among adolescents, with the critical age of peak drug use occurring among young people between the ages of 18 and 25 [4,5]. This is due to the strong tendency to experiment, curiosity, susceptibility to peer pressure, rebellion against authority, and low self-esteem that make such individuals vulnerable to drug use and abuse [6]. Substance abuse impairs the successful transition to adulthood by interfering with the development of critical thinking and the learning of important cognitive skills [7]. Higher rates of physical and mental illness and poorer overall health and well-being are also reported among adolescents who abuse drugs [8]. Reports had acknowledged the awareness of such consequences among college students [3,7].

Some of the risk factors associated with substance use include the presence of early mental and behavioral health problems, peer pressure, poorly equipped schools, poverty, poor parental supervision and relationships, poor family structure, lack of opportunity, isolation, gender, and access to drugs. Protective factors include high self-esteem, religiosity, grit, peer factors, self-control, parental monitoring, academic competence, anti-drug policies, and strong neighborhood ties [2,9–13]. With regard to alcohol consumption, environmental factors such as economic development, culture, availability of alcohol and the level and effectiveness of alcohol policies are relevant factors in explaining differences and historical trends in alcohol consumption and related harm [14].

The rapid economic, social, and cultural transitions that most countries in sub-Saharan Africa are now experiencing, have created conditions conducive to increased and socially disruptive use of drugs and alcohol [2].

Reports from Cameroun indicate high prevalence of substance use among university students [3,12]. Data on risk factors related to substance use among this specific group are scarce, justifying the conduct of this survey whose objective was to depict determinants related to alcohol, smoking and drug use among university students in Yaoundé, the capital of Cameroon.

## Methods

### Study design & period

An institutional descriptive and cross-sectional study was conducted at Yaounde 1 University in Cameroon from September to October 2023.

### Setting

Yaoundé is the political capital of the Central Region of Cameroon. The population is estimated at 1.5 million and includes all of the country’s ethnic groups. The demographic structure is characterized by a very young population (people under the age of twenty make up 60% of the population) [15]. The University of Yaoundé I is a non-profit public institution of higher education located in the metropolis of Yaoundé. It educates between 10,000 and 15,000 students each year. The university has many faculties and professional schools, the most important of which are in the fields of literature, basic sciences, medicine and education.

### Study participants & selection criteria

All students attending classes at the University of Yaounde during the study period were eligible. All eligible students aged 18 years and older from the selected university institution, regardless of their year of study, who gave written informed consent were included.

### Sampling method

A convenience non probabilistic technique was used to enroll consenting students.

### Data collection tool and procedures

The study tool was a questionnaire designed to collect sociodemographic information and data on smoking habits (cigarettes, water pipes), alcohol and drug use (marijuana, cocaine, tramadol, heroin, etc.). Data were collected during an interview conducted by trained investigators. Prior to enrolment, potential participants were informed of the aims and objectives of the study before written informed consent was obtained.

### Data quality assurance

Data collectors were medical students who received 2 days of training and instruction prior to the survey. The data collected were reviewed and checked for completeness prior to data entry.

### Data processing and analysis

The predictor consisted of sociodemographic characteristics including age, gender, level of study, living situation, faculty/school, living on campus, district of residence, residential area, social charge, drug use, and smoking. Four dependent variables were assessed, including alcohol use, smoking, drug use, and polysubstance use. Polysubstance use refers to the use of more than one substance in close temporal proximity with overlapping effects. [16–18]. Data were checked, entered, recoded as necessary, and analyzed using R Statistics version 4.2.3. Fisher’s exact test was used to compare proportions. Simple and multiple binary logistic regressions were used to assess the strength of association between variables. The selection of predictors that best fit the model was done stepwise using the Akaike Information Criterion (AIC) [19]. The model with the lowest index was then selected. A p-value <0.05 was considered statistically significant.

### Ethical Approval Statement

This study has received the authorization of the Faculty of Medicine and Biomedical Sciences (N°: 472/UY/FMBS). Informed consent was obtained from participants prior to inclusion in the study. All methods were performed in accordance with declaration of Helsinki.

## Results

### Socio-demographic Profile

A total of 191 university students were enrolled in the study. The median age was 20 years. They were mostly male (66.5%) and aged 20-25. Students living with the family (67.5%) or living outside (72.8%) the campus were the most represented (Table 1).

**Table 1.** Factors Associated with Alcohol Use among University Students in Yaounde, 2023 (*n=*191)

| Predictor | Alcohol Use<br>*n* (%) |  | COR | *p*-value | AOR | 95% CI |  | *p*-value |
| --- | --- | --- | --- | --- | --- | --- | --- | --- |
|  | Yes<br>*n*=126 | No<br>*n*=65 |  |  |  | Lower | Upper |  |
| **Sex** |  |  |  |  |  |  |  |  |
| Female | 42 (65.6) | 22 (34.4) | 1 |  |  |  |  |  |
| Male | 84 (66.1) | 43 (33.9) | 1.02 | 0.943 |  |  |  |  |
| **Age group** |  |  |  |  |  |  |  |  |
| <20 | 36 (50.7) | 35 (49.3) | 1 |  | 1 | — | — |  |
| 20-25 | 65 (74.7) | 22 (25.3) | 2.87 | **0.002** | 2.89 | 1.45 | 5.91 | **0.003** |
| 25-30 | 17 (73.9) | 6 (26.1) | 2.75 | 0.056 | 2.37 | 0.85 | 7.37 | 0.112 |
| 30-43 | 8 (80.0) | 2 (20.0) | 3.89 | 0.099 | 4.72 | 1.06 | 33.3 | 0.063 |
| **Living situation** |  |  |  |  |  |  |  |  |
| Family | 83 (64.3) | 46 (35.7) | 1 |  |  |  |  |  |
| Alone | 31 (64.6) | 17 (35.4) | 1.01 | 0.976 |  |  |  |  |
| Collocation | 12 (85.7) | 2 (14.3) | 3.33 | 0.126 |  |  |  |  |
| **Study level** |  |  |  |  |  |  |  |  |
| Bachelor | 98 (63.2) | 57 (36.8) | 1 |  |  |  |  |  |
| Master | 24 (82.8) | 5 (17.2) | 2.79 | **0.048** |  |  |  |  |
| Doctorate | 4 (57.1) | 3 (42.9) | 0.78 | 0.745 |  |  |  |  |
| **Faculty & school** |  |  |  |  |  |  |  |  |
| Literature | 18 (72.0) | 7 (28.0) | 1 |  |  |  |  |  |
| Sciences | 82 (64.6) | 45 (35.4) | 0.71 | 0.475 |  |  |  |  |
| Medicine | 16 (64.0) | 9 (36.0) | 0.69 | 0.545 |  |  |  |  |
| Polytechnic | 6 (66.7) | 3 (33.3) | 0.78 | 0.764 |  |  |  |  |
| Others | 4 (80.0) | 1 (20.0) | 1.56 | 0.713 |  |  |  |  |
| **Living in the campus** |  |  |  |  |  |  |  |  |
| Yes | 30 (57.7) | 22 (42.3) | 1 |  | 1 | — | — |  |
| No | 96 (69.1) | 43 (30.9) | 1.64 | 0.141 | 1.72 | 0.84 | 3.50 | 0.134 |
| **District of residence** |  |  |  |  |  |  |  |  |
| Nkolndongo | 4 (57.1) | 3 (42.9) | 1 |  |  |  |  |  |
| Biyem-Assi | 23 (59.0) | 16 (41.0) | 1.08 | 0.928 |  |  |  |  |
| Djoungolo | 7 (87.5) | 1 (12.5) | 5.25 | 0.207 |  |  |  |  |
| Efoulan | 66 (66.0) | 34 (34.0) | 1.46 | 0.635 |  |  |  |  |
| Mvog-Ada | 4 (66.7) | 2 (33.3) | 1.50 | 0.725 |  |  |  |  |
| Nkolbisson | 8 (80.0) | 2 (20.0) | 3.00 | 0.318 |  |  |  |  |
| Odza | 6 (75.0) | 2 (25.0) | 2.25 | 0.468 |  |  |  |  |
| Others | 8 (61.5) | 5 (38.5) | 1.20 | 0.848 |  |  |  |  |
| **Residential area** |  |  |  |  |  |  |  |  |
| Urban | 113 (65.7) | 59 (34.3) | 1 |  |  |  |  |  |
| Rural | 13 (68.4) | 6 (31.6) | 1.13 | 0.812 |  |  |  |  |
| **Social charge** |  |  |  |  |  |  |  |  |
| Yes | 19 (70.4) | 8 (29.6) |  |  |  |  |  |  |
| No | 107 (65.2) | 57 (34.8) | 0.79 | 0.603 |  |  |  |  |
| **Smoking** |  |  |  |  |  |  |  |  |
| No | 75 (59.1) | 52 (40.9) | 1 |  | 1 | — | — |  |
| Yes | 51 (79.7) | 13 (20.3) | 2.72 | **0.005** | 2.68 | 1.31 | 5.72 | **0.008** |
| **Drug use** |  |  |  |  |  |  |  |  |
| No | 87 (62.1) | 53 (37.9) | 1 |  |  |  |  |  |
| Yes | 39 (76.5) | 12 (23.5) | 1.98 | 0.067 |  |  |  |  |

COR: Crude Odds Ratio; AOR: Adjusted Odds Ratio; CI: Confidence Interval

### Alcohol Use

Age (*p*-value=0.002), level of study (*p*-value=0.048), and smoking status (*p*-value=0.005) of the participants were significant factors associated with alcohol on univariate analysis. However, multivariate logistic regression showed that students aged 20-25 years were significantly 2.9 times more likely to drink alcohol than those aged less than 20 years (*p*-value=0.003). Students who smoke were 2.7 times more likely to drink alcohol than those who do not smoke (*p*-value=0.008) (Table 1).

### Smoking

Living situation (*p*-value=0.013) and drug use status (*p*-value<0.0001) were significant factors associated with smoking on univariate analysis. On multivariate analysis, drug users were 3.2 times more likely to smoke than drug non-users (*p*-value<0.0001) (Table 2).

**Table 2.**
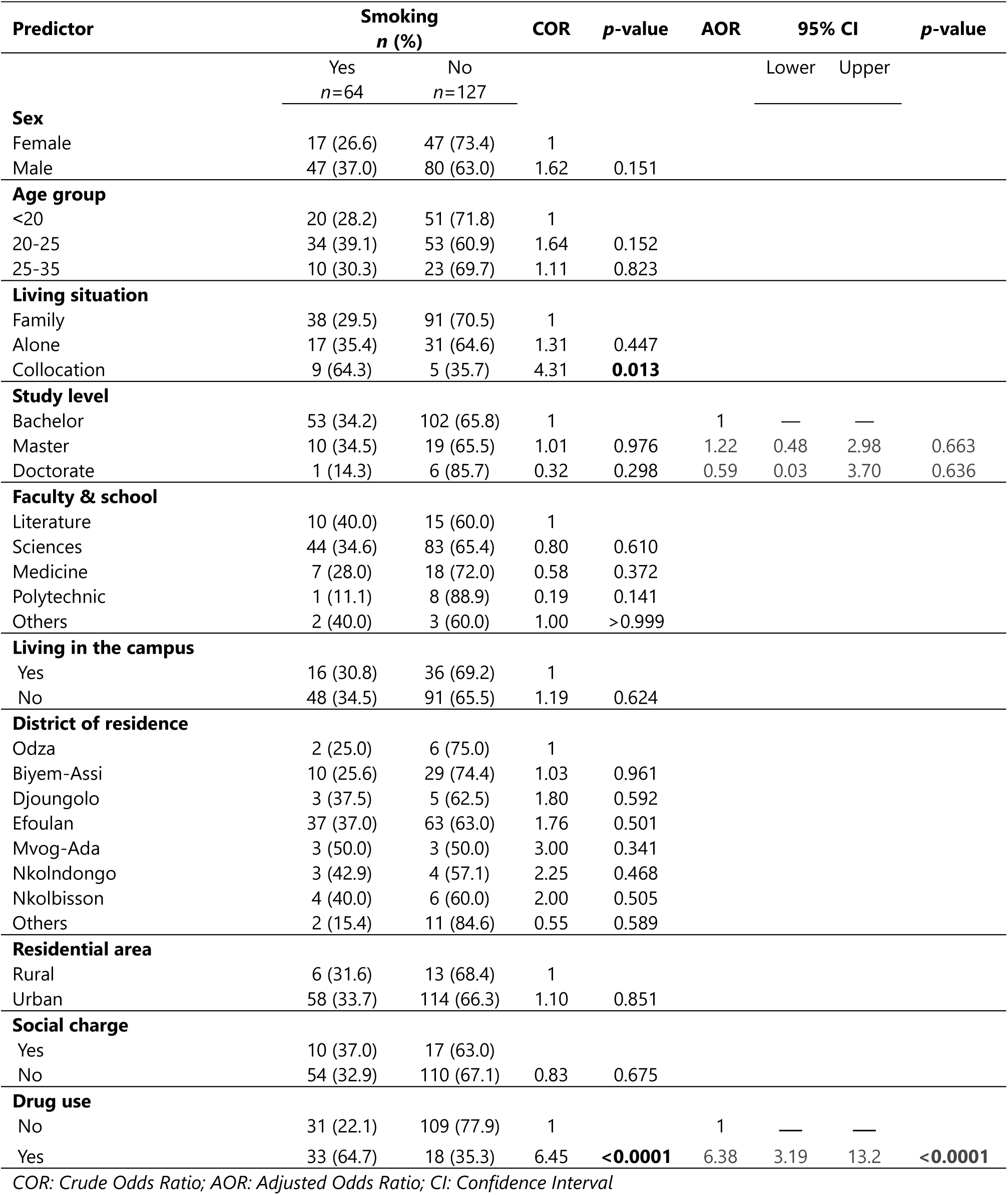
Predictors of Smoking among University Students in Yaounde, 2023 (*n=*191)

| Predictor | Smoking<br>*n* (%) |  | COR | *p*-value | AOR | 95% CI |  | *p*-value |
| --- | --- | --- | --- | --- | --- | --- | --- | --- |
|  | Yes<br>*n*=64 | No<br>*n*=127 |  |  |  | Lower | Upper |  |
| **Sex** |  |  |  |  |  |  |  |  |
| Female | 17 (26.6) | 47 (73.4) | 1 |  |  |  |  |  |
| Male | 47 (37.0) | 80 (63.0) | 1.62 | 0.151 |  |  |  |  |
| **Age group** |  |  |  |  |  |  |  |  |
| <20 | 20 (28.2) | 51 (71.8) | 1 |  |  |  |  |  |
| 20-25 | 34 (39.1) | 53 (60.9) | 1.64 | 0.152 |  |  |  |  |
| 25-35 | 10 (30.3) | 23 (69.7) | 1.11 | 0.823 |  |  |  |  |
| **Living situation** |  |  |  |  |  |  |  |  |
| Family | 38 (29.5) | 91 (70.5) | 1 |  |  |  |  |  |
| Alone | 17 (35.4) | 31 (64.6) | 1.31 | 0.447 |  |  |  |  |
| Collocation | 9 (64.3) | 5 (35.7) | 4.31 | **0.013** |  |  |  |  |
| **Study level** |  |  |  |  |  |  |  |  |
| Bachelor | 53 (34.2) | 102 (65.8) | 1 |  | 1 | — | — |  |
| Master | 10 (34.5) | 19 (65.5) | 1.01 | 0.976 | 1.22 | 0.48 | 2.98 | 0.663 |
| Doctorate | 1 (14.3) | 6 (85.7) | 0.32 | 0.298 | 0.59 | 0.03 | 3.70 | 0.636 |
| **Faculty & school** |  |  |  |  |  |  |  |  |
| Literature | 10 (40.0) | 15 (60.0) | 1 |  |  |  |  |  |
| Sciences | 44 (34.6) | 83 (65.4) | 0.80 | 0.610 |  |  |  |  |
| Medicine | 7 (28.0) | 18 (72.0) | 0.58 | 0.372 |  |  |  |  |
| Polytechnic | 1 (11.1) | 8 (88.9) | 0.19 | 0.141 |  |  |  |  |
| Others | 2 (40.0) | 3 (60.0) | 1.00 | >0.999 |  |  |  |  |
| **Living in the campus** |  |  |  |  |  |  |  |  |
| Yes | 16 (30.8) | 36 (69.2) | 1 |  |  |  |  |  |
| No | 48 (34.5) | 91 (65.5) | 1.19 | 0.624 |  |  |  |  |
| **District of residence** |  |  |  |  |  |  |  |  |
| Odza | 2 (25.0) | 6 (75.0) | 1 |  |  |  |  |  |
| Biyem-Assi | 10 (25.6) | 29 (74.4) | 1.03 | 0.961 |  |  |  |  |
| Djoungolo | 3 (37.5) | 5 (62.5) | 1.80 | 0.592 |  |  |  |  |
| Efoulan | 37 (37.0) | 63 (63.0) | 1.76 | 0.501 |  |  |  |  |
| Mvog-Ada | 3 (50.0) | 3 (50.0) | 3.00 | 0.341 |  |  |  |  |
| Nkolndongo | 3 (42.9) | 4 (57.1) | 2.25 | 0.468 |  |  |  |  |
| Nkolbisson | 4 (40.0) | 6 (60.0) | 2.00 | 0.505 |  |  |  |  |
| Others | 2 (15.4) | 11 (84.6) | 0.55 | 0.589 |  |  |  |  |
| **Residential area** |  |  |  |  |  |  |  |  |
| Rural | 6 (31.6) | 13 (68.4) | 1 |  |  |  |  |  |
| Urban | 58 (33.7) | 114 (66.3) | 1.10 | 0.851 |  |  |  |  |
| **Social charge** |  |  |  |  |  |  |  |  |
| Yes | 10 (37.0) | 17 (63.0) |  |  |  |  |  |  |
| No | 54 (32.9) | 110 (67.1) | 0.83 | 0.675 |  |  |  |  |
| **Drug use** |  |  |  |  |  |  |  |  |
| No | 31 (22.1) | 109 (77.9) | 1 |  | 1 | — | — |  |
| Yes | 33 (64.7) | 18 (35.3) | 6.45 | **<0.0001** | 6.38 | 3.19 | 13.2 | **<0.0001** |

COR: Crude Odds Ratio; AOR: Adjusted Odds Ratio; CI: Confidence Interval

### Drug Use

Drug use was significantly associated with the district of residence of the user on univariate analysis. The resident of Nkolndongo seems to be more prone to drug use compared to students from Biyem-Assi district (p-value=0.024) (Table 3).

**Table 3.** Factors Associated with Drug Use among University Students in Yaounde, 2023 (*n=*191)

| Predictor | Drug Use<br>*n* (%) |  | COR | *p*-value | AOR | 95% CI |  | *p*-value |
| --- | --- | --- | --- | --- | --- | --- | --- | --- |
|  | Yes<br>*n*=51 | No<br>*n*=140 |  |  |  | Lower | Upper |  |
| **Sex** |  |  |  |  |  |  |  |  |
| Female | 13 (20.3) | 51 (79.7) | 1 |  |  |  |  |  |
| Male | 38 (29.9) | 89 (70.1) | 1.68 | 0.151 |  |  |  |  |
| **Age group** |  |  |  |  |  |  |  |  |
| <20 | 20 (28.2) | 51 (71.8) |  |  |  |  |  |  |
| 20-25 | 22 (25.3) | 65 (74.7) | 0.86 | 0.683 |  |  |  |  |
| 25-30 | 8 (34.8) | 15 (65.2) | 1.36 | 0.547 |  |  |  |  |
| 30-43 | 1 (10.0) | 9 (90.0) | 0.28 | 0.246 |  |  |  |  |
| **Living situation** |  |  |  |  |  |  |  |  |
| Family | 32 (24.8) | 97 (75.2) | 1 |  |  |  |  |  |
| Alone | 12 (25.0) | 36 (75.0) | 1.01 | 0.979 |  |  |  |  |
| Collocation | 7 (50.0) | 7 (50.0) | 3.03 | 0.053 |  |  |  |  |
| **Study level** |  |  |  |  |  |  |  |  |
| Bachelor | 45 (29.0) | 110 (71.0) | 1 |  | 1 | — | — |  |
| Master | 6 (20.7) | 23 (79.3) | 0.64 | 0.360 | 0.64 | 0.22 | 1.58 | 0.360 |
| Doctorate | 0 (0.0) | 7 (100.0) | 0.00 | 0.986 |  |  |  |  |
| **Faculty & school** |  |  |  |  |  |  |  |  |
| Literature | 10 (40.0) | 15 (60.0) | 1 |  |  |  |  |  |
| Sciences | 32 (25.2) | 95 (74.8) | 0.51 | 0.135 |  |  |  |  |
| Medicine | 7 (28.0) | 18 (72.0) | 0.58 | 0.372 |  |  |  |  |
| Polytechnic | 1 (11.1) | 8 (88.9) | 0.19 | 0.141 |  |  |  |  |
| Others | 1 (20.0) | 4 (80.0) | 0.38 | 0.410 |  |  |  |  |
| **Living in the campus** |  |  |  |  |  |  |  |  |
| Yes | 14 (26.9) | 38 (73.1) | 1 |  |  |  |  |  |
| No | 37 (26.6) | 102 (73.4) | 0.98 | 0.966 |  |  |  |  |
| **District of residence** |  |  |  |  |  |  |  |  |
| Biyem-Assi | 6 (15.4) | 33 (84.6) | 1 |  |  |  |  |  |
| Odza | 2 (25.0) | 6 (75.0) | 1.83 | 0.514 |  |  |  |  |
| Djoungolo | 3 (37.5) | 5 (62.5) | 3.30 | 0.162 |  |  |  |  |
| Efoulan | 31 (31.0) | 69 (69.0) | 2.47 | 0.067 |  |  |  |  |
| Mvog-Ada | 1 (16.7) | 5 (83.3) | 1.10 | 0.936 |  |  |  |  |
| Nkolndongo | 4 (57.1) | 3 (42.9) | 7.33 | **0.024** |  |  |  |  |
| Nkolbisson | 2 (20.0) | 8 (80.0) | 1.38 | 0.725 |  |  |  |  |
| Others | 2 (15.4) | 11 (84.6) | 1.00 | 0.999 |  |  |  |  |
| **Residential area** |  |  |  |  |  |  |  |  |
| Rural | 4 (21.1) | 15 (78.9) | 1 |  |  |  |  |  |
| Urban | 47 (27.3) | 125 (72.7) | 1.41 | 0.402 |  |  |  |  |
| **Social charge** |  |  |  |  |  |  |  |  |
| Yes | 9 (33.3) | 18 (66.7) | 1 |  |  |  |  |  |
| No | 42 (25.6) | 122 (74.4) | 0.69 | 0.4024 |  |  |  |  |

COR: Crude Odds Ratio; AOR: Adjusted Odds Ratio; CI: Confidence Interval

### Polysubstance Use

Living situation (*p*-value=0.016), faculty/school(*p*-value=0.04), and district of residence (*p*-value=0.037) were significantly associated with polysubstance use. Students living in collocation were 3.8 times more likely to be use more than two substances regularly than those living with their families (*p*-value=0.023) (Table 4).

**Table 4.** Predictors of Polysubstance Use among University Students in Yaoundé, 2023 (*n=*191)

| Predictor | Polysubstance Use<br>*n* (%) |  | COR | *p*-value | AOR | 95% CI |  | *p*-value |
| --- | --- | --- | --- | --- | --- | --- | --- | --- |
|  | Yes<br>*n*=67 | No<br>*n*=124 |  |  |  | Lower | Upper |  |
| **Sex** |  |  |  |  |  |  |  |  |
| Female | 17 (26.6) | 47 (73.4) | — |  |  |  |  |  |
| Male | 50 (39.4) | 77 (60.6) | 1.80 | 0,082 |  |  |  |  |
| **Age group** |  |  |  |  |  |  |  |  |
| <20 | 20 (28.2) | 51 (71.8) |  |  |  |  |  |  |
| 20-25 | 36 (41.4) | 51 (58.6) | 1.80 | 0,086 |  |  |  |  |
| 25-43 | 11 (33.3) | 22 (66.7) | 1.28 | 0,592 |  |  |  |  |
| **Living situation** |  |  |  |  |  |  |  |  |
| Family | 39 (30.2) | 90 (69.8) | — |  | — | — | — |  |
| Alone | 19 (39.6) | 29 (60.4) | 1.51 | 0,240 | 1.33 | 0.63 | 2.78 | 0.448 |
| Collocation | 9 (64.3) | 5 (35.7) | 4.15 | **0,016** | 3.81 | 1.23 | 13.1 | **0.023** |
| **Study level** |  |  |  |  |  |  |  |  |
| Bachelor | 54 (34.8) | 101 (65.2) | — |  | — | — | — |  |
| Master | 13 (44.8) | 16 (55.2) | 0.64 | 0,307 | 1.39 | 0.58 | 3.29 | 0.456 |
| Doctorate | 0 (0.0) | 7 (100.0) | 0.00 | 0,986 |  |  |  |  |
| **Faculty & school** |  |  |  |  |  |  |  |  |
| Literature | 14 (56.0) | 11 (44.0) | — |  |  |  |  |  |
| Sciences | 43 (33.9) | 84 (66.1) | 0.40 | **0,040** |  |  |  |  |
| Medicine | 7 (28.0) | 18 (72.0) | 0.31 | **0,048** |  |  |  |  |
| Polytechnic | 1 (11.1) | 8 (88.9) | 0.10 | **0,041** |  |  |  |  |
| Others | 2 (40.0) | 3 (60.0) | 0.52 | 0,517 |  |  |  |  |
| **Living in the campus** |  |  |  |  |  |  |  |  |
| Yes | 18 (34.6) | 34 (65.4) | — |  |  |  |  |  |
| No | 49 (35.3) | 90 (64.7) | 1.03 | 0.935 |  |  |  |  |
| **District of residence** |  |  |  |  |  |  |  |  |
| Biyem-Assi | 9 (23.1) | 30 (76.9) | — |  |  |  |  |  |
| Odza | 2 (25.0) | 6 (75.0) | 1.11 | 0,907 |  |  |  |  |
| Djoungolo | 5 (62.5) | 3 (37.5) | 5.56 | **0,037** |  |  |  |  |
| Efoulan | 40 (40.0) | 60 (60.0) | 2.22 | 0,064 |  |  |  |  |
| Mvog-Ada | 2 (33.3) | 4 (66.7) | 1.67 | 0,589 |  |  |  |  |
| Nkolndongo | 3 (42.9) | 4 (57.1) | 2.50 | 0,283 |  |  |  |  |
| Nkolbisson | 4 (40.0) | 6 (60.0) | 2.22 | 0,286 |  |  |  |  |
| Others | 2 (15.4) | 11 (84.6) | 0.61 | 0,559 |  |  |  |  |
| **Residential area** |  |  |  |  |  |  |  |  |
| Rural | 6 (31.6) | 13 (68.4) | — |  |  |  |  |  |
| Urban | 61 (35.5) | 111 (64.5) | 1.19 | 0,736 |  |  |  |  |
| **Social charge** |  |  |  |  |  |  |  |  |
| Yes | 12 (44.4) | 15 (55.6) | — |  |  |  |  |  |
| No | 55 (33.5%) | 109 (66.5) | 0.63 | 0,274 |  |  |  |  |

COR: Crude Odds Ratio; AOR: Adjusted Odds Ratio; CI: Confidence Interval

Most substance users were aware not only of the academic consequences but also of the psychosocial, psychological, and health consequences of substance use. In this regard, almost all smokers (95.1%) reported being aware the of the psychosocial, mental and health consequences of substance use (*p*-value=0.021) (Table 5).

**Table 5.** Perception of some Substances Uses Consequences among University Students in Yaounde, 2023 (*n=*191)

| Feel the need to stop | Academic consequence<br>*n* (%) |  | Total (100%) | *p*-value<sup>1</sup> |
| --- | --- | --- | --- | --- |
|  | No<br>*n*=23 | Yes<br>*n*=103 |  |  |
| **Alcohol use** |  |  |  | 0.351 |
| No | 11 (15.1) | 62 (84.9) | 73 |  |
| Yes | 12 (22.6) | 41 (77.4) | 53 |  |
| **Smoking** |  |  |  | 0.178 |
| No | 11 (47.8) | 12 (52.2) | 23 |  |
| Yes | 12 (29.3) | 29 (70.7) | 41 |  |
| **Drug use** |  |  |  | 0.272 |
| No | 5 (35.7) | 9 (64.3) | 14 |  |
| Yes | 7 (18.9) | 30 (81.1) | 37 |  |
| **Psychosocial, mental and health consequence** |  |  |  |  |
|  | *n* (%) |  |  |  |
|  | No<br>*n*=10 | Yes<br>*n*=116 |  |  |
| **Alcohol use** |  |  |  | >0.999 |
| No | 6 (8.2) | 67 (91.8) | 73 |  |
| Yes | 4 (7.5) | 49 (92.5) | 53 |  |
| **Smoking** |  |  |  | 0,021 |
| No | 6 (26.1) | 17 (73.9) | 23 |  |
| Yes | 2 (4.9) | 39 (95.1) | 41 |  |
| **Drug use** |  |  |  | 0.478 |
| No | 1 (7.1) | 13 (92.9) | 14 |  |
| Yes | 1 (2.7) | 36 (97.3) | 37 |  |

<sup>1</sup>Fisher exact test

## Discussion

Multivariate logistic regression showed that students aged 20-25 years were significantly 2.9 times more likely to drink alcohol than those aged less than 20 years. This particular age group may be influenced by the university environment, including peer pressure. A study reports that college students use alcohol to relieve stress and improve mood [20]. This result corroborates findings in Myanmar and a previous study in Cameroon, where student age was a key factor associated with alcohol consumption, with older students more likely to consume alcohol [12,13]. Students who smoke were 2.7 times more likely to drink alcohol than those who do not smoke. According to epidemiological research, alcohol consumption and high rates of smoking among adults and adolescents are strongly correlated; in fact, the rate of smoking among alcoholics is thought to be at least twice that of the general population [21–23]. This combination of alcohol and tobacco smoke has serious health consequences, including an increased risk of certain diseases and a higher risk of death from stroke and cancer [21,23]. However, this association appears to be stronger at higher levels of consumption: heavy drinkers are more likely to smoke heavily, and vice versa [24]. Our result confirms observations in Turkey and Myanmar [10,13].

Regarding the smoking status of college students, those who used drugs were 3.2 times more likely to smoke than those who did not use drugs. Often referred to as “gateway drugs”, tobacco and alcohol are among the first substances used by young people. Their easy accessibility is likely a contributing factor, in addition to other biological and social factors (e.g., peer pressure, acculturation, family history of substance use problems, etc.) [22,25,26]. Study results show that, like alcohol, cigarette smoking among adolescents is strongly associated with illicit drug use. In addition to more frequent use of illicit drugs, adolescents who smoke consistently throughout adolescence are at significantly greater risk for abuse or dependence on marijuana and other drugs [22]. Our findings are consistent with those observed in Germany and the USA [27,28].

Drug use was significantly associated with the user’s District of residence on univariate analysis. The resident of Nkolndongo seems to be more prone to drug use compared to students residing in Biyem-Assi District. This may be explained by the social environment of this particular District of Yaoundé, where anecdotal evidence describes a context in which the drug market in the area is prolific and the promotion of alcohol consumption is facilitated by the multiplicity of bars, even in the vicinity of educational institutions [29,30]. Almost all studies reported substantial evidence on individual risk variables for adolescent substance dependence, which can be divided into five subdomains, including personal/individual characteristics, significant adverse childhood experiences, personal mental health diagnoses, drug history, comorbidity, and individual attitudes and perspectives [5]. This suggests that further research should be conducted in the hotspot area of Yaoundé to gain a better understanding of this social scourge.

Polysubstance use, which refers to the use of more than one substance at the same time, is increasingly recognized as a pressing public health issue [16–18]. In our study, students living in shared housing were 3.8 times more likely to be polysubstance users than those living with family. Peer motivation and lack of parental control may have facilitated the use of many substances at once [3,31]. Most substance users were aware not only of their academic consequences but also of their psychosocial, mental and health consequences. Our findings corroborate findings among college students in Ethiopia, where most of the negative effects attributed by respondents to substance use were related to substance use [32].

## Limitations

The statistical inference of these results should take into account that the study included only students from one tertiary institution and used a non-probability sampling technique. However, the study provides insight and specific sociodemographic determinants that drive the expansion and sustainability of the phenomenon among university students.

## Conclusions

This study has identified several significant risks related to substance use. Such determinants included age, living situation, District of residence, smoking and drug use status. Substance abuse among university students requires special attention, urgent preventive measures and targeted information, education and communication efforts. This determinant should help to tailor specific policies and strategies, including health education initiatives, to explain the long-term consequences of substance use. In addition, additional qualitative analyses should be conducted to better understand the underlying causes of behaviors that lead to the use of alcohol, tobacco and other illicit substances.

## Data Availability

All data produced in the present work are contained in the manuscript

https://biomedgrid.com/fulltext/volume21/prevalence-of-substance-use-and-related-%20behaviors-among-tertiary-students-a-cross-sectional-survey-in-yaound%C3%A9-cameroon.002812.php

## Declarations

### Authors’ Contribution

Study conception & design: FZLC; Data collection: FZLC and MFE; Data analysis and interpretation: FZLC; Drafting of original manuscript: FZLC; Critical revision of the manuscript: FZLC, MFE, AA and TM; Final approval of the manuscript: All authors.

### Consent for publication

Not applicable.

### Availability of data and materials

All data generated or analyzed during this study are included in this published article.

## Acknowledgements

Our gratitude goes to undergraduate students of the faculty of medicine who gave a support in the conduction of the study.

